# Compression of motor unit recruitment threshold patterns are present in the subacute phase post stroke

**DOI:** 10.1101/2025.03.31.25324516

**Authors:** Mio Ito, Takanori Ito, Hayase Funakoshi, Kei Takahata, Nina Suresh, Takanori Kokubun

**Author notes:** Correspondence: Takanori Kokubun Ph.D. Graduate School of Health, Medicine, and Welfare, Saitama Prefectural University, Department of Physical Therapy, School of Health and Social Services, Saitama Prefectural University, 820 San-nomiya, Koshigaya-Shi, Saitama, 343-8540, Japan.

## Abstract

Voluntary contraction anomalies of post-stroke survivors progress from flaccid paralysis to recovery of upper extremity motor function in the subacute phase. However, muscle weakness often persists, and it is unclear what changes or aberrations persist in neuromuscular function, particularly in motor unit behavior. Our objective was to characterize motor unit discharge behavior in hemiplegic stroke patients in the subacute phase. We tested seven subacute stroke patients at two time points (Timepoint 1 and Timepoint 2) a minimum of two weeks apart during the subacute phase. We used wireless surface electromyography to detect motor unit activities on both sides of our tested participants. Participants carried out two types of target force tracking tasks with isometric elbow flexion. We performed 2-way ANOVA between the time point and test side. The recruitment threshold force(RTF) of the Ramp task exhibited a significant interaction between the Timepoint and Test side (*p* < 0.00). The post hoc test showed the RTF of the affected side was not significantly lower than the contralateral side (*p* = 0.99) at Timepoint 1. On the other hand, the affected side at Timepoint 2 was significantly lower than the contralateral side (*p* < 0.00). The low recruitment threshold on the affected side may be more exacerbated than the contralateral side chronologically during the subacute phase of stroke. Our results suggest that the assessment of motor units in the subacute phase of stroke can contribute to the early detection of abnormal neuromuscular activity and, thereby, the establishment of effective rehabilitation.

**NEW & NOTEWORTHY:** This study clarified altered chronological motor unit recruitment patterns in the subacute stroke. We revealed the neuromuscular physiological abnormalities on the affected side may persist from the subacute period to the chronic stage. To maximize recovery of motor function in stroke patients with prolonged symptoms, it is necessary to detect neuromuscular dysfunction in the subacute phase and establish early prevention. This study provided fundamental knowledge on preventive rehabilitation of persistent paresis during the subacute phase. Keywords: stroke; motor unit; surface electromyography; subacute; rehabilitation

## INTRODUCTION

Impairment of voluntary motor function is one of the most prevalent issues in stroke survivors. In the sequelae of stroke, voluntary muscle contraction disorders, such as paresis or voluntary muscle weakness (1), directly impact activities of daily living. In particular, motor dysfunction of the upper limb occurs early after stroke onset and greatly affects patients’ activities of daily living. (2, 3) Paresis in post-stroke shows differences in the degree of motor function chronologically from the sub-acute phase to the chronic phase. The subacute phase is defined as 7 days to 6 month after onset, while the chronic phase is defined as 6 months after onset from stroke. (4) In particular, it has been reported that the subacute phase shows a clear recovery (5), and this recovery trend is reported regardless of the lesion location or phases from stroke onset (6, 7). However, the neurophysiological recovery mechanisms from voluntary contraction disorders in post-stroke remain unclear. In a healthy state, the efferent pathway of voluntary muscle contraction includes a complicated neural system: cerebral activity, corticospinal tract, neuro-muscular junction, and muscle contraction. In stroke patients, every component has the possibility of changing from the onset to the subacute phase; however, most research has focused on cerebral activity as the central pathophysiological root (8).

Recently, analysis of motor unit behavior is critical in the assessment of mechanisms of impaired voluntary muscle contraction. Motor units are the smallest functional units of voluntary muscle activation, and motor unit analysis is the only known method to characterize spinal neural control of muscle in human subjects. (9) A motor unit consists of one motor neuron and its innervated muscle fibers. Analyzing the activity of motor units as physiological structures provides an essential understanding of underlying impairments in neural control in voluntary muscle weakness. This is because the assessment of motor unit activation in subacute stroke survivors can provide information regarding whether muscle weakness is partially due to deficits in neural control and potential changes in passive muscle properties.

It is known that motor unit activities of paralyzed muscles in chronic stroke survivors showed abnormal recruitment patterns. Previous studies of chronic stroke survivors showed a decrease in motor unit discharge rate (10-17), and in general a compression of the force recruitment patterns of motor units was reported (13, 18, 19). These studies evaluated muscles such as tibialis anterior, biceps brachii, first dorsal interosseous, triceps, peroneus longus, and vastus lateralis. Although the severity of motor dysfunction after stroke has been shown to alter chronologically, motor function recovery is most promoted only in the subacute phase. This means the motor function of stroke patients will probably show positive and negative changes from 1 to 6 months after the onset of stroke, affected by many factors: portion and degree of cerebrum damage, rehabilitation therapy, and individual physical activity level. Although we know that motor function is restored the subacute phase, how the neuromuscular properties of recovery are unknown specifically motor unit recruitment and discharge rates.

The objective of this study was to characterize and quantify motor unit recruitment patterns during a target tracking task in hemiplegic stroke patients of upper extremity, specifically the biceps brachii muscle during the subacute phase. Our hypothesis is that if the process of motor function of subacute stroke patients shows recovery, the neuromuscular physiological condition will be improved with increased recruitment and rate cording patterns of motor units. The clarification of motor unit recruitment patterns through the recovery process of patients with moderate paralysis provides fundamental insights into the neurophysiological changes that underlie the recovery mechanism and can lead to an effective rehabilitation strategy.

## MATERIALS AND METHODS

### 1. Participants

We recruited seven stroke patients admitted to a rehabilitation hospital and obtained written informed consent for this study (Table 1). The first set of experimental measurements (Timepoint 1) was performed within 1-3 months after stroke onset, and each participant was tested again after the first set of measurements (Timepoint 2) to observe chronological changes. The accurate range between timepoints as 19-38 days and that Timepoint 1 was at 1-4 months.

**Table 1.**
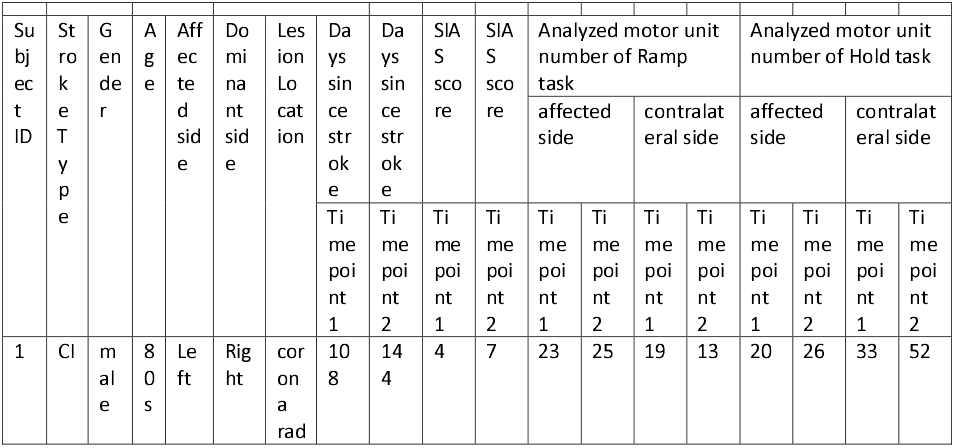
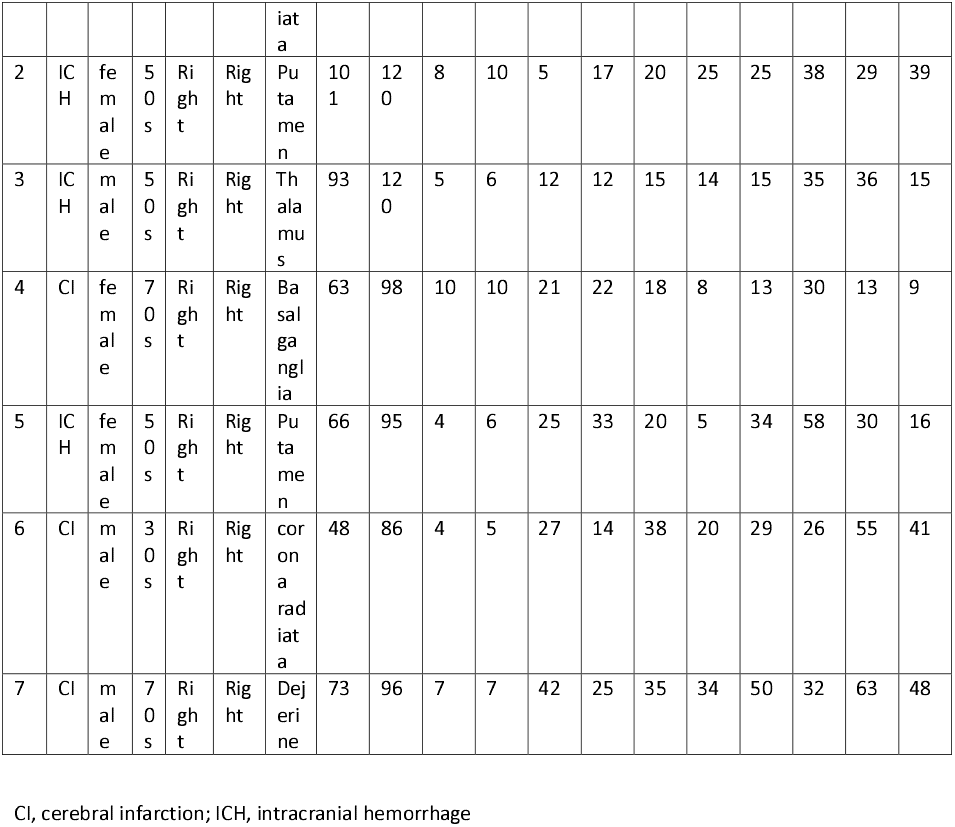
Subject characteristics for the 7 stroke patients who participated in the experiments

We included participants with initial hemispheric stroke onset who have hemiplegia and excluded participants with severe upper extremity joint range of motion limitation, cognitive dysfunction, and higher cerebrum dysfunction to the extent that the experiment cannot be carried out. Rehabilitation Amakusa Hospital’s ethics review committee approved this study (Approval number: 20210202).

### 2. Experimental setup

We used wireless surface electromyography (sEMG: Trigno Galileo sensor, Delsys. Inc) to detect motor unit activities. Each sensor has four channels of sEMG signals (Figure 1 - A). We placed the wireless sEMG electrodes on each participant’s affected and contralateral biceps brachii. The electrodes were placed to the line between the medial acromion and the fossa cubit at 1/3 from the fossa cubit based on the SENIAM guidelines. To place the electrodes in the same location, we measured the distance from the medial acromion to the electrode placement position on the line from the medial acromion to the fossa cubit. The position of the electrode at Timepoint 2 was determined based on this distance. Additionally, we recorded forces using a one-degrees-of-freedom load cell (Model SSM-AJ-250, Interface. Inc) with force feedback provided to the participant on a portable computer monitor. We connected a load cell to the original experimental device, which was made according to the previous study (20). The participants were seated with the elbow joint flexes 90 degrees and the shoulder joint abducted flexed 90degrees, and the distal forearm and load cell were fixed. Then, the PC monitor was placed in front of the participant to show the target line in the task of exerting isometric elbow flexion force (Figure 1 - B). Force analog data recorded from the loadcell was sampled at 1,926Hz after it was filtered by bandwidth: DC-50Hz. EMG works software (Delsys. Inc) synchronized sEMG signals, the force signal from a load cell projected on the monitor.

**Figure 1.**
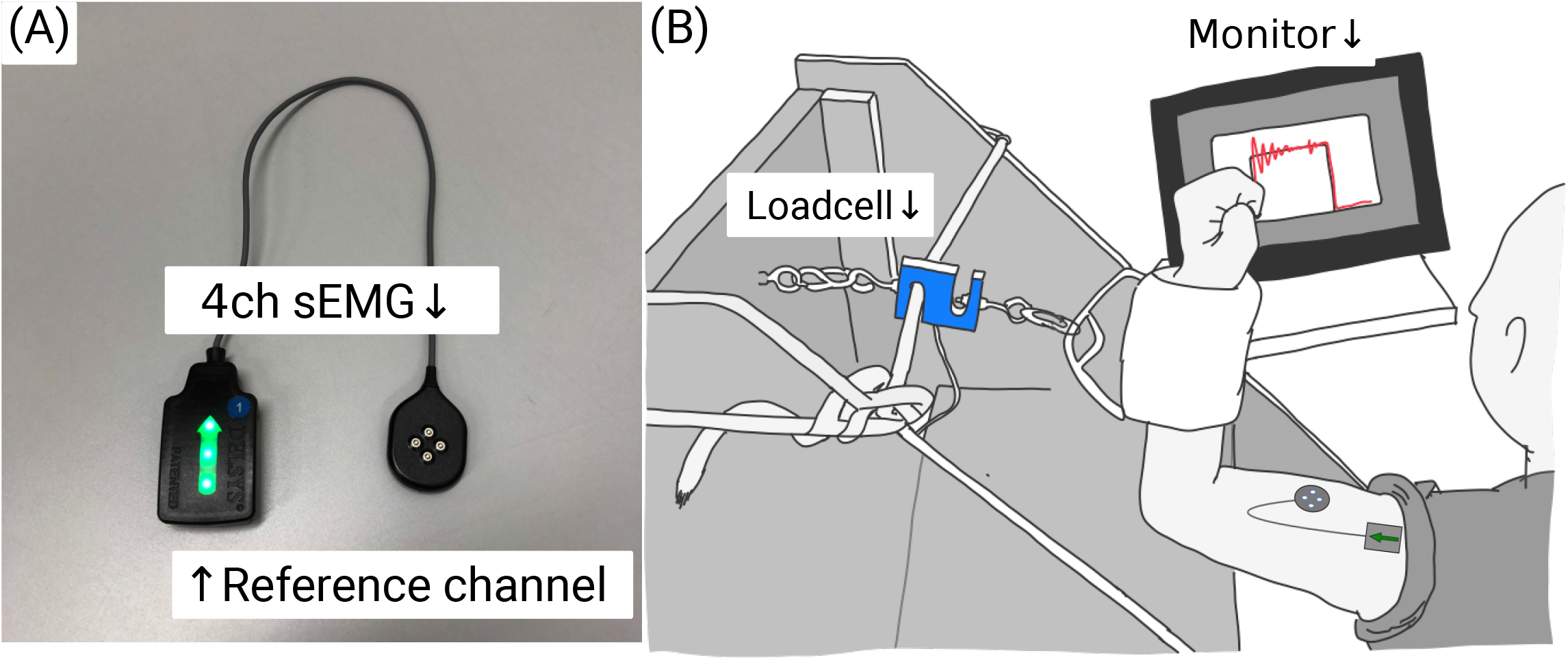
Experimental set up A: 4ch sEMG electrodes to detect motor unit activities. B: We connected between a load cell and participant’s forearm.

### 3. Experimental Procedures

First, participants carried out the elbow flexion maximum voluntary contraction (MVC) task. The participants performed the MVC task while checking the feedback of their force level on the monitor. The larger value of the two trials was designated as the MVC. Next, participants performed two types of target force tracking tasks. One was the Hold task, maintaining constant force generation for 15 seconds with elbow flexion isometric contractility at 80%MVC. Another was the Ramp task, which gradually increased the force to 80%MVC over 15 seconds at a speed of 5.3%MVC/sec and maintained at 80%MVC over 3 seconds. Each participant performed the MVC task twice and the target force tracking task three times on both the affected and contralateral sides. Participants took a 5-minute rest between each trial.

### 4. Data processing of sEMG signals and motor unit

sEMG was sampled at 2,222Hz, and bandpass filter was 20-450Hz. Motor unit action potentials were decomposed from the recorded sEMG signals using the Neuromap software algorithm (Delsys. Inc). We used the time series data of the timing of detected motor unit spikes as the motor unit action potential train (Figure 2). We analyzed motor units with an estimation accuracy of at least 80 percent (the software provided these statistics) and used the dataset with five or more motor units detected (17).

**Figure 2.**
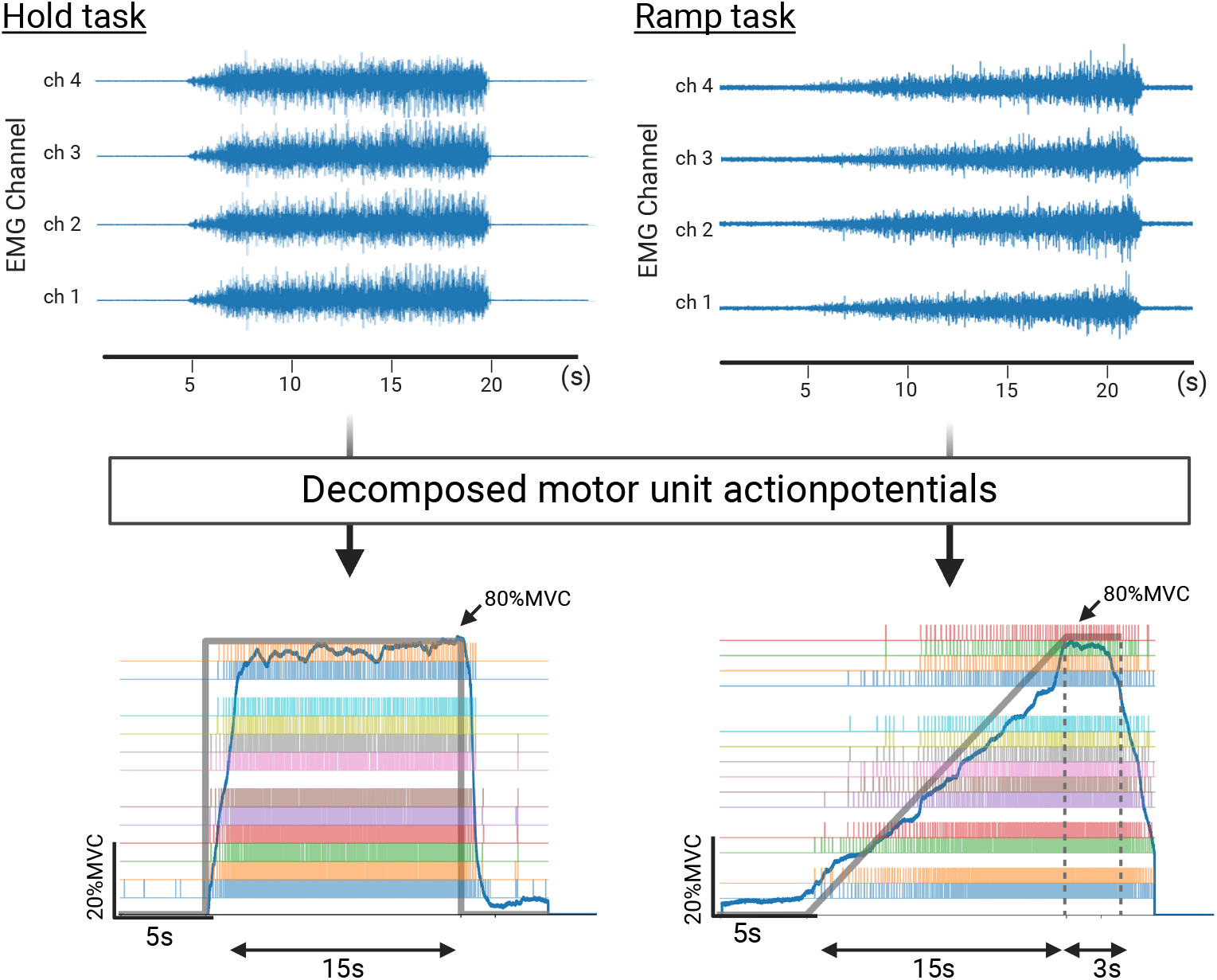
Data processing of sEMG signals and motor unit Surface EMG signals of Hold task and Ramp task. We decomposed motor unit action potentials by Neuromap system. We plotted colorfully bar plots indicate motor units firing, respectively. The gray line shows the target line and the blue line shows the force exerted by the subject.

### 5. Maximum muscle contraction force and force tracking accuracy

We computed the largest of the two trials of MVC Force and the root means square error during both target force tracking tasks (RMSE of Hold task, RMSE of Ramp task). MVC Force was the maximum value of the force of maximum isometric contraction of elbow flexion during the MVC task. The RMSE of the measured values relative to the target line indicated the accuracy of force regulation during each target force tracking task. The representative value was the mean value of the three trials of RMSE. We used RMSE to evaluate the accuracy of reproducing the participant’s Task.

### 6. Motor unit recruitment and discharge rate estimation

For the motor unit data, we report the number of decomposed motor units used in the analysis, recruitment threshold force, and discharge rate of each decomposed motor unit. We calculated the number of decomposed motor units as the total number of motor units extracted in Hold tasks and Ramp tasks. The recruitment threshold force was calculated as the %MVC Force at the time the MU started to discharge (21) with an inter-spike interval < 200ms (14) in the Ramp task. We included only the Recruitment threshold from 5 seconds after a five-second silent period. The mean discharge rate at the Hold task was calculated from a 15s averaging window during the 80%MVC force steadiness (Figure 3 - A). The peak discharge rate of the Ramp task was calculated at the end of the ramp force generation; a 3-s window was used because the force was relatively constant and the discharge rate was relatively stable (Figure 3 - B). (12)

**Figure 3.**
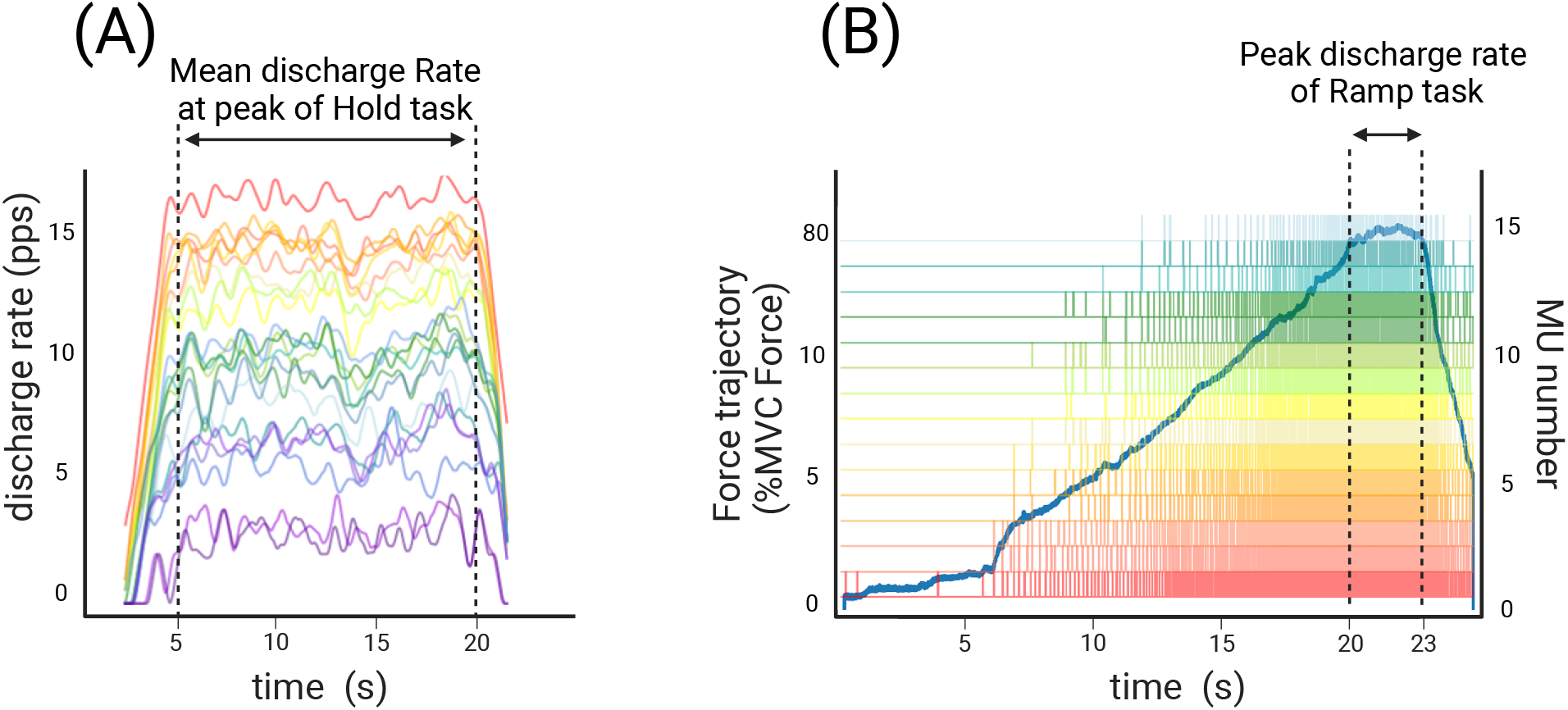
Exemplar discharge rate trace and discharge timing plot A: Each color trace represents the discharge rate over time of one exemplar trial of Hold task. Different colors represent different motor units. The mean discharge rate of a 15-s window as marked by the dashed lines at Hold task was calculated as the mean discharge rate. PPS represents pulses per second. B: Each color bar plot represents the discharge timing of one exemplar trial of Ramp task. The peak discharge rate of a 3-s window as marked by the dashed lines at the end of Ramp task was calculated as the peak discharge rate. The blue trace represents the force output. %MVC Force represents percentage normalized MVC Force. Motor unit (MU) number represents detected number of motor units.

### 7. Clinical assessment score of post-stroke motor function

To assess motor function on the affected side, we also evaluated the SIAS (Stroke Impairment Assessment Set) upper extremity score as the clinical score. It was evaluated on a 10-point scale for the upper extremity. In this study, we assessed the upper extremity motor function using a sum of the “Motor function” items (SIAS score).

### 8. Statistical analysis

We performed a two-way analysis of variance (2way ANOVA) using Timepoint (Timepoint 1 or Timepoint 2) and Test side (the affected side or the contralateral side) as factors after checking for normality. 2way ANOVA was performed on MVC force values, Force RMSE, Mean discharge rate of Hold task, Recruitment threshold force during Ramp task, Peak discharge rate during Ramp task. We used Tukey’s post hoc tests to identify the location of statistical differences when appropriate. We performed a paired t-test on the SIAS score after checking for normality at each time point. *p* < 0.05 was considered statistically significant.

## RESULTS

The total number of motor units analyzed in the Ramp task, at Timepoint 1 on the affected side was 155, and the contralateral side was 165. At Timepoint 2 on the affected side was 148, and the contralateral side was 119. The total number of motor units analyzed in the Hold task, at Timepoint 1 on the affected side was 186, and the contralateral side was 259. At Timepoint 2 on the affected side was 245, and the contralateral side was 220.

The results of the 2-way ANOVA are shown in Table 2. Each value of the results is listed as mean ± standard deviation.

**Table 2.**
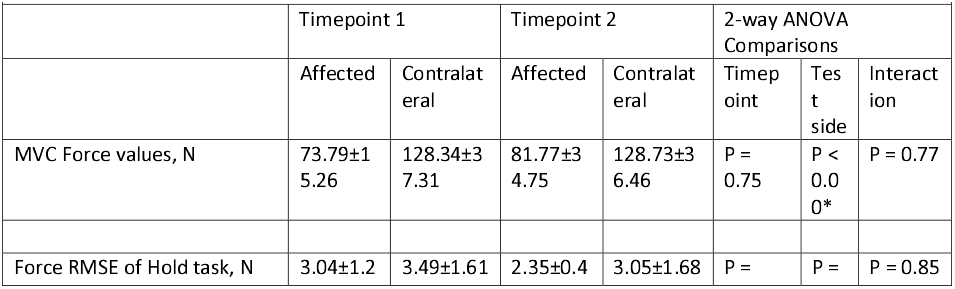
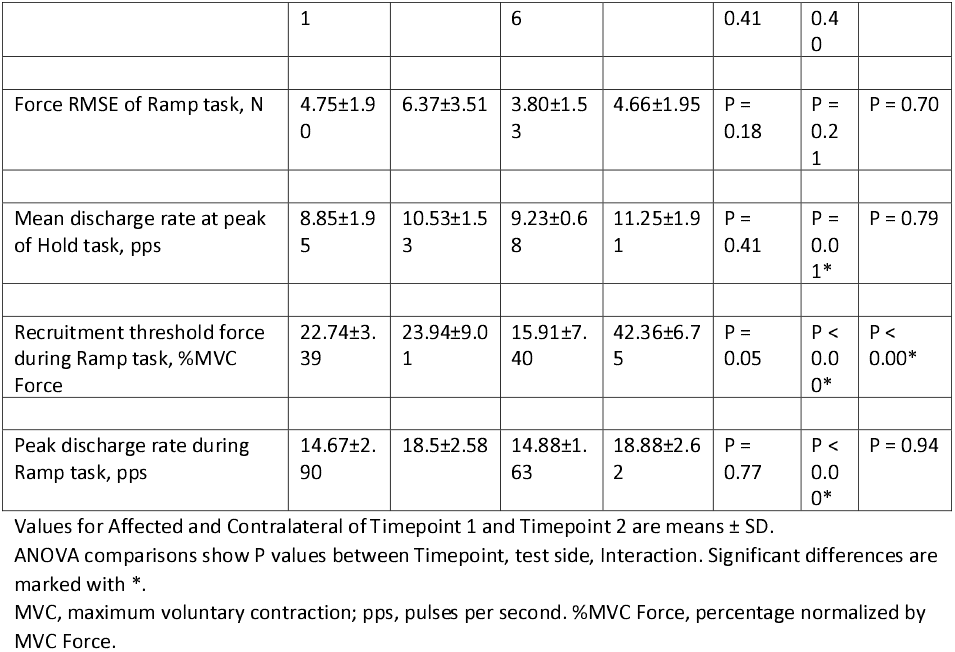
2way ANOVA of force and motor unit characteristics

The recruitment threshold force during Ramp task at Timepoint 1 on the affected side was 22.74±3.39 %MVC, and on the contralateral side was 23.94±9.01 %MVC. The recruitment threshold force for Timepoint 2 on the affected side was 15.91±7.40 %MVC, and on the contralateral side was 42.36±6.75 %MVC. There was a significant difference for the Test side (*p* < 0.00), while there was no significance of the Timepoint (*p* = 0.05). There was significant interaction between the Timepoint and Test side (*p* < 0.00).

Results of the post hoc test of the Recruitment threshold force during Ramp task are shown in Figure 4-A. The Tukey-Kramer results, conducted as a post hoc test, showed the contralateral side of Timepoint 1 was significantly lower than the contralateral side of Timepoint 2 (*p* < 0.00). On the other hand, the affected side of Timepoint 1 was not significantly different from the affected side of Timepoint 2 (*p* = 0.34). At Timepoint 1, the affected side was not significantly lower than the contralateral side (*p* = 0.99). On the other hand, at Timepoint 2, the affected side was significantly lower than the contralateral side (*p* < 0.00).

**Figure 4.**
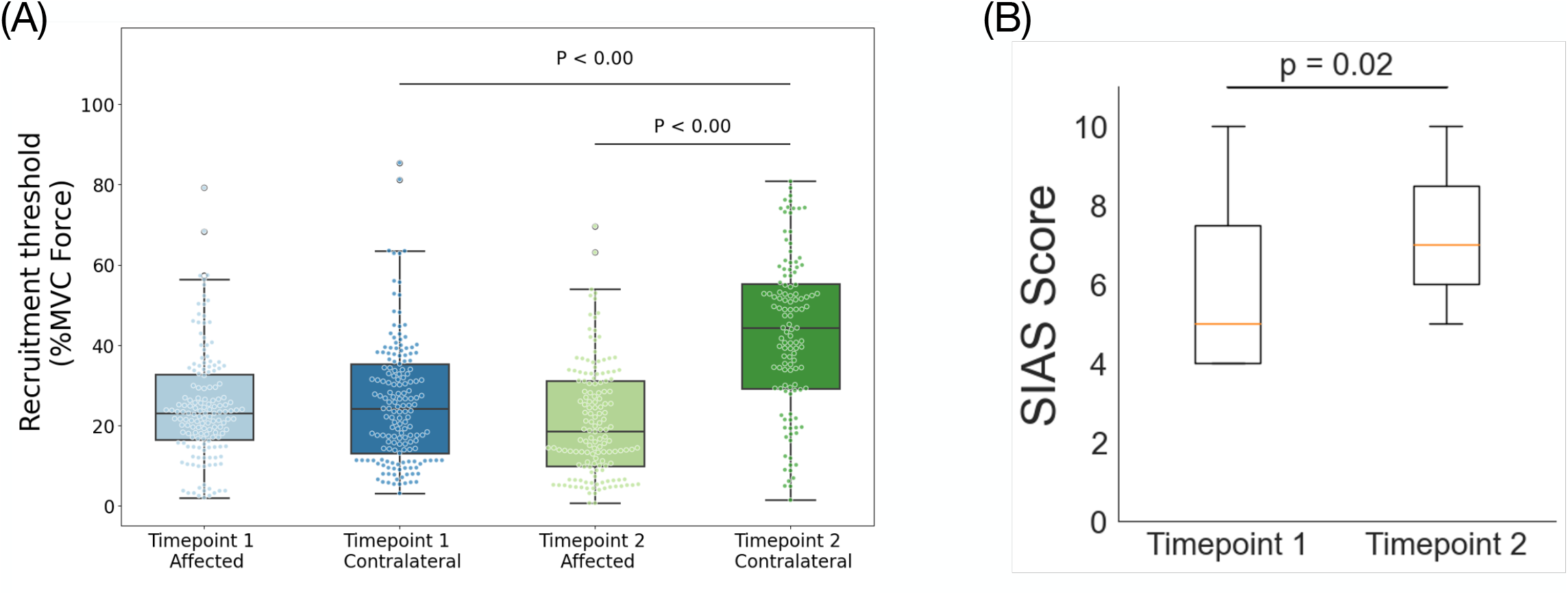
Results of post hoc test of Recruitment threshold force during Ramp task and results of t test of SIAS score A: Results of post hoc test of Recruitment threshold in Ramp task. B: results of t test of SIAS score

The mean discharge rate in Timepoint 1 on the affected side was 8.85±1.95 pps, and on the contralateral side was 10.53±1.53 pps. The mean discharge rate at Timepoint 2 on the affected side was 9.23±0.68 pps, and on the contralateral side was 11.25±1.91 pps. There was no significance for the

Timepoint (*p* = 0.41), although a significant difference was found for the Test side (*p* = 0.01). There was no significant interaction between the Timepoint and Test side (*p* = 0.79).

The peak discharge rate at the Ramp task in Timepoint 1 on the affected side was 14.67±2.90 pps, and on the contralateral side was 18.5±2.58 pps. For timepoint 2, the peak discharge rate on the affected side was 14.88±1.63 pps, and on the contralateral side was 18.88±2.62 pps. There was no significance for Timepoint (*p* = 0.77), while there were significant differences on the Test side (*p* < 0.00). There was no significant interaction between the Timepoint and Test side (*p* = 0.94).

The result of the SIAS score, which indicates the clinical motor function of stroke is shown in Figure 4-B. SIAS score at Timepoint 1 was 6.0±2.20, and at Timepoint 2 was 7.28±1.83. SIAS score at Timepoint 2 was significantly higher than the SIAS score at Timepoint 1 (*p* = 0.02).

The MVC force values at Timepoint 1 on the affected side was 73.79±15.26 N, and on the contralateral side was 128.34±37.31 N, for Timepoint 2 on the affected side was 81.77±34.75 N, and the contralateral side was 128.73±36.46 N. The MVC force on the affected side was statistically lower than the contralateral side at both time points. There was no significance for the Timepoint (*p* = 0.75), although a significant difference was found for the Test side (*p* < 0.00). There was no significant interaction between the Timepoint and Test side (*p* = 0.77).

Regarding the Force RMSE of the Hold task, the RMSE at Timepoint 1 on the affected side was 3.04±1.21 N, and on the contralateral side, it was 3.49±1.61 N. The RMSE at Timepoint 2 on the affected side was 2.35±0.46 N, and on the contralateral side was 3.05±1.68 N. There was no significance regarding either the Timepoint (*p* = 0.41) or Test side (*p* = 0.40). There was also no significant interaction between the Timepoint and Test side (*p* = 0.85). In the RMSE of the Ramp task, the RMSE at Timepoint 1 on the affected side was 4.75±1.90 N, and on the contralateral side was 6.37±3.51 N. The RMSE at Timepoint 2 on the affected side was 3.80±1.53 N and on the contralateral side was 4.66±1.95 N. There was no significance regarding either the Timepoint (*p* = 0.18) or the Test side (*p* = 0.21). There was also no significant interaction between the Timepoint and Test side (*p* = 0.70).

## DISCUSSION

To analyze the chronological characteristics of impaired motor function subsequent to a hemispheric stroke of upper extremity, many researchers have been focused on the central nervous system, assessing cortical activity over a lengthy time period. In contrast, this study focused on neuro-muscular function, in particular, motor unit activities, to characterize spinal motor pathways in voluntary movement. In this study, we report on clinical, force, and motor unit data collected from both sides of a post-stroke cohort at two different time points during the subacute phase. Contrary to our hypothesis, motor units did not change despite the improvement in clinical motor function.

The Recruitment threshold force during Ramp task was significantly higher in Timepoint 2 than in Timepoint 1 on the contralateral side, while the affected side had no significant differences. In other words, the contralateral side showed significant improvement over time, but the recruitment threshold on the affected side remains compressed. Such a recruitment pattern that deflects to the lower recruitment threshold in a force ramp-up task is called compression of the recruitment threshold. It has been reported as a characteristic neuromuscular dysfunction in stroke patients, and it may be a direct result of muscle atrophy and inefficient activation of motor units (13, 18, 19). In muscle which compression of the recruitment threshold is observed, fatigable motor units are recruited early. As a result, the recruited motor units are overloaded and are unable to generate forces. (13) Therefore, this abnormal neuromuscular physiological characteristic adversely affects the muscle.

A unique feature of these results was that Timepoint 1 showed the same compression of the recruitment threshold on the contralateral side as on the affected side. Bilateral motor dysfunction in stroke has been reported at the neuromuscular functional level, including motor units (15, 22, 23). The compression of the recruitment threshold observed in Timepoint 1 in this study showed a significant improvement trend in Timepoint 2 one month later. Therefore, it is suggested that abnormal neuromuscular physiological features occur bilaterally in stroke, but the recovery process may differ between the affected and the contralateral sides.

On the affected side, compression of the recruitment threshold was commonly observed in Timepoint 1 and Timepoint 2. When the corticospinal tract was damaged by stroke, motor commands from the primary motor cortex to the spinal cord (24)were reduced. As reported in previous studies (18), it is likely that on the affected side, motor units with low thresholds were selectively preferentially recruited, but those with high thresholds were not. When motor units with these lower thresholds are selectively recruited, the synchronous firing of these motor units puts a strain on certain motor units. Moreover, if these abnormal recruitment patterns of motor units are reinforced at the subacute phase, the risk is increased for atrophy of type II fibers, which are governed by motor units with high firing thresholds. Compression of the recruitment threshold mentioned in previous studies has been reported in studies of chronic stroke patients more than one year after stroke onset (13). In this study, we included the earlier stages of stroke onset, and the same tendency was commonly observed at the two-time points in the recovery period. Therefore, it is suggested that the compression of the recruitment threshold may be an abnormal motor unit recruitment pattern that remains long-term effect from subacute phase to chronic phase after the onset of stroke.

To investigate the motor unit rate coding patterns, participants performed the Hold task and Ramp task. Both the mean discharge rate of Hold task and peak discharge rate during Ramp task were no significant effects of the timepoint factor while significant effect of the test side factor, with the affected side being lower than the contralateral side, our study results are consistent with those of the chronic stroke study (14, 15). This indicates that low discharge rates on the affected side may begin early and can be a long-term potential motor control issue after stroke.

We discuss the chronological relationship between the results of the clinical motor function assessment scale and the motor unit data of the affected side. Although the SIAS score significantly improved from Timepoint 1 to Timepoint 2, the Recruitment threshold force during Ramp task, Mean discharge rate at the peak of the Hold task, and Peak discharge rate of the Ramp task did not change significantly from Timepoint 1 and Timepoint 2. This suggests that there was a gap between the results of existing chronological clinical assessments of motor function and physiological data representing the neuromuscular physiological characteristics of the affected muscle. Correlation between severity of motor function and motor unit data has been reported in chronic stroke (14). As shown in this study, negative results of potential muscles from the subacute phase may impair the motor function of stroke over time. Therefore, the function of the affected side in the subacute phase should not be captured based on one aspect of the existing motor function assessment. It may prolong neuromuscular physiological abnormalities due to chronological muscle degeneration in stroke patients.

The SIAS score used in this study was the same as that used in a previous study (25), with two items on a 5-point scale representing upper limb motor function on the affected side. Most of the existing motor function assessment scales for the affected side do not reflect the potential neuromuscular physiological characteristics since they are mainly scored according to whether voluntary movements are possible or not. Previous studies have expressed concern about the aspects of these motor functions of clinical scores and have argued for the establishment of indices that represent true recovery (26).

The results of this study indicate that the degree of motor dysfunction evaluated clinically only evaluates whether voluntary movement is possible or not among the motor functions of the affected side. It may not be an indicator that can evaluate the true recovery of the affected side. Therefore, we argue that the assessment of voluntary movements should be separated from the assessment of muscle parenchyma. We suggest that assessment of clinical motor function alone does not capture the true physiological abnormalities latent in the affected side. In the future, when assessing the function of the affected side of stroke patients, if we can assess the activity of motor units in addition to exiting assessment indices, we may be able to detect neuromuscular physiological abnormalities that cannot be detected by clinical motor function assessment.

In summary, these results represent the difference in chronological motor unit characteristics of the affected side and the contralateral sides in the subacute phase of stroke. These results indicate that even at the subacute stage, there are differences between the affected and contralateral sides. However, there are no differences between the tested two time points. Our results suggest that the neurophysiological aberrations occur early in the acute stage post-stroke and remain relatively constant thereafter. The results also indicated a gap between clinical rating scales of motor dysfunction and neurophysiological characteristics, as the clinical ratings show improvements, whereas the experimental parameters, which have greater precision and resolution, exhibit no changes over time in the subacute phase post-stroke. Detailed neuromuscular assessment may be necessary in the subacute phase, when motor function is dynamically changing. These findings suggest that analysis of the motor unit activity in subacute patients after stroke is beneficial to improve stroke recovery for rehabilitation intervention.

This study has some limitations. First, the degree of hemiplegia of the participant is only moderate and not covered in a wide range of patients. The data of sEMG was essential to detect motor unit activities. However, detecting sEMG in participants with hemiplegia was not easy, so the EMG signal could not be stable and had to be excluded in case of noisy data. Therefore, we focused on moderate stroke patients for whom we could detect their EMG signal. Consequently, the knowledge we revealed in this study only applied to understanding the neurophysiological characteristics of moderate stroke hemiplegics. Second, the number of participants included in the analysis is small. Currently, the method of decomposing motor units from sEMG has not yet been widely applied to stroke patients, especially in the subacute phase. So, we tried to investigate the motor unit recruitment patterns using sEMG in subacute stroke patients. The selection of participants in this study is variously constrained by the fact that subacute stroke patients are hospitalized. The number of participants in this study was determined by referring to previous studies investigating motor unit activity (12). The exclusion criteria we have established are sufficient to reflect the neurophysiological information of humans correctly. Third, rehabilitation interventions for the included patients were not limited. However, all patients in the study received a similar three hours of rehabilitation per day at the same hospital, which did not significantly affect the results. Furthermore, this study was not able to verify whether the same motor units were present across time points. It is important to compare the same motor unit of a subject between two time points in order to determine how the characteristics of the motor unit changed over time. In the future study, the motor unit tracking technique could be utilized to present more robust data.

## Conclusion

This study clarified the altered chronological motor unit recruitment patterns in the subacute phase after stroke. The results revealed that the neuromuscular physiological abnormalities in the affected side may persist from Subacute to long term. In addition, the existing clinical motor function assessment used in the subacute phase may not detect actual physiological abnormalities on the affected side. To maximize recovery of motor function in stroke patients with prolonged symptoms, it is necessary to detect neuromuscular dysfunction in the subacute phase and establish early prevention. This study provided basic knowledge on preventive rehabilitation interventions during the subacute phase.

## Supporting information

Table 1. Subject characteristics for the 7 stroke patients who participated in the experiments

Table 2. Result of 2way ANOVA

## DATA AVAILABILITY

The data that support the findings of this study are available from the corresponding author upon reasonable request.

## ACKNOWLEDGMENTS

The graphical abstracts, Figures 1, 2, and 3 were created with BioRender.com.

## DISCLAIMERS

We have no disclaimers.

## AUTHOR CONTRIBUTIONS

M.I. and T.K. conceived and designed research; performed experiments; analyzed data; interpreted results of experiments; prepared figures; drafted manuscript; edited and revised manuscript; and approved final version of the manuscript.

T.I. and H.F. supported the analysis of the data.

T.I., H.F., K.T., and N.S. interpreted the results of experiments, edited and revised the manuscript, and approved the final version of the manuscript.

